# Pre-Diagnosis Observational and Prescription History Associated with Alzheimer’s Disease Incidence

**DOI:** 10.1101/2024.07.17.24310553

**Authors:** Gareth Williams

## Abstract

**Background:** Alzheimer’s disease (AD) has so far proved refractory to intervention. However, disease incidence is variable across prior medication and observational measures. The present study adopted a data-driven approach to inform possible drug repurposing strategies in the light of concurrent prescription and biometric data.

**Methods and Results:** A real-world dataset was harnessed to compare observational and prescription data for 250,000 individuals prior to AD diagnosis against an age-, sex-, and clinical practice-matched control cohort with no AD history. Observation data was shown to both explain the associations of classes of drug prescription with AD incidence and to bolster the repurposing potential of the GLP-1 agonist class of anti-diabetic drugs.

**Conclusion:** This study provides insights into how drug repurposing based on prescription histories can be informed by concurrent observational data. These findings offer novel insights to be explored in future research on causal models for AD progression.

## Introduction

With the exception of the controversial approval for AD of the at best moderately effective anti-amyloid antibody therapies[1–4], with aducanumab being recently discontinued due to cost and side effect issues[5], there have been no new AD therapeutics coming to market in over 20 years. The inherently complex nature of the disease with multiple pathological features associated with various biological pathways and risk factors encompassing lifestyle[6] and genetic variants[7] has led researchers to go so far as to question AD’s status as a disease and argue that it should be rather considered a syndrome[8] due to the mismatch in causal factors and ultimate presentation. Turning away from target-based drug discovery approaches, the wealth of data on the disease has informed lifestyle modifications[9, 10] and motivated a search for repurposing candidates where existing approved therapeutics with extensive safety data and prescription histories are hypothesised as candidate therapeutics for diseases for which they weren’t initially developed[11, 12]. Other neurodegenerative conditions like Parkinson’s disease (PD) have also been the subject of repurposing efforts[13]. Repurposing can take many forms from the emergence of targets shared with other diseases[14] to the correlation of high content biological data, such as gene expression profiles[15, 16], between disease and compound. A more direct approach is based on epidemiology, where low disease incidence associations with drug prescription may inform a new intervention route, as shown with the emergence of salbutamol as a potential protective against PD though an analysis of the Norwegian NorPD prescription database [17]. However, without a viable biological mechanism, epidemiology-based associations do not establish causality but rather inform future research through generating hypotheses.

The COVID pandemic precipitated the Health Data Research UK (HDRUK), through the British Heart Foundation Data Science Centre, making available primary health care data for 57 million people in England to researchers in the hope of gaining insights into amelioration strategies[18]. The HDRUK data is currently available for COVID pandemic related studies, at least on a fee free basis. However, on a smaller scale, the well-established Clinical Practice Research Datalink (CPRD)[19, 20] provides curated data on approximately 19 million currently contributing (67 million historical) patient’s observation and prescription histories from across the UK. CPRD provides an ideal platform to investigate associations between disease incidence and prior prescription history and observational data and has been the basis of investigating associations of dementia with inflammatory biomarkers[21], comorbidities like type 2 diabetes (T2D) [22] and drug use[23, 24]. Other initiatives like the UK Biobank[25] provide the researcher with valuable diagnostic data linked to genetic variant information and tissue samples for over half a million individuals.

In the present work we sought to harness electronic health records from a large real-world dataset to investigate the associations, both positive and negative, between AD incidence and prior prescription history and observational data. Specifically, we collected a cohort with an AD diagnosis and defined the index date as the date of first diagnosis. Everyone with an AD diagnosis was matched with people of the same age, sex and clinical practice with no AD diagnosis at index. This effectively constituted a retrospective case-control study to investigate disease risk factors prior to onset. We found that high systolic blood pressure (SBP) and low blood albumin levels are significantly associated with a higher incidence of AD up to ten years prior to diagnosis. A combination of blood-borne biomarkers and biometric data gathered 10 to 20 years prior to diagnosis of AD facilitated a modest predictability of future AD incidence with an operating characteristic (ROC) area under the curve (AUC) of 0.79+/-0.001. An analysis of prescription histories revealed both positive and negative associations of drug prescription with AD incidence. An analysis of concurrent observational data was used to investigate the possible causal relationships between drug use and AD incidence. Notably, the negative association of anti-hypertensives with AD incidence was explained by SBP being higher in the cohort going on to develop AD. The highly significant negative association of the migraine triptan medications was shown to be confounded by both SBP levels and migraine incidence. Two classes of T2D medications were shown to be associated with a lower AD incidence and this bolstered by the observation of a higher AD incidence in the T2D sub cohort. Our analysis within the female cohort recapitulates the reported AD risk lowering potential of hormonal therapies. It is hoped that this analysis will help build hypotheses around new repurposing candidates and lend support to existing AD therapy candidates in trials.

## Methods

### Cohort

The present study used routinely collected data from the CPRD, one of the world largest primary care electronic health records datasets. The CPRD contains medical records for about 24% of the UK primary care population with complete prescribing data and clinical diagnosis information, laboratory tests, and referrals made following a primary care consultation. The CPRD population is representative in terms of size, patients’ socio-demographic attributes, and geographical distribution of practices. The data have been validated extensively for pharmaco-epidemiological, clinical and public health investigations. The study population included patients aged 50 years or over at the time of a first diagnosis of AD between 1^st^ January 2012 and 31^st^ December 2022. These patients were individually matched on age, sex and practice with a set of controls without AD, see Table 1. There were on average six controls per AD individual. AD diagnosis was defined using SNOMED CT and Read medical codes reflective of an AD event (see Supplementary Table S1 for a complete list of codes). Data was extracted from the CPRD Aurum version in September 2023.

**Table 1.** The case-control cohort characteristics for AD.

|  | WOMEN |  | MEN |  |
| --- | --- | --- | --- | --- |
|  | AGE(mean+/-sd) | N | AGE(mean+/-sd) | N |
| AD | 83.22+/-7.96 | 157195 | 80.88+/-8.11 | 103709 |
| CTL | 83.22+/-7.97 | 931257 | 80.88+/-8.10 | 616870 |

### Observations

Individuals were assigned to T2D and migraine status based on observations corresponding to the SNOMED CT codes in Supplementary Tables S2 and S3. The observational biometric and blood borne variables defined for the cohort are listed in Supplementary Table S4. Each measure has a mean and standard deviation and measures outside 2 standard deviations of the mean were dropped as erroneous or outliers. The measures have various coverage over the cohorts, see Supplementary Table S4.

### Medication

Medication incidence is determined by prescription events for any of the British National Formulary (BNF) (www-medicinescomplete-com.apollo.worc.ac.uk/#/browse/bnf) (1364) medications, all prescription variants of a given BNF type are pooled. Two different time ranges prior to index were analysed, pooling prescription data 5 to 10 and 10 to 20 years prior diagnosis. To address the differential coverage of prescription data the cohort matching was refined so that the overall medication frequency or observation frequency over the given age range prior to index period was non-zero and within 20% for AD and non-AD pairing. The cohort characteristics 5 to 10 and 10 to 20 years prior to index are shown in Table 2.

**Table 2.** The cohort summary for the medication association with AD incidence analysis. Cases were matched with controls based on age, sex, clinical practice together with the number of observations and prescriptions over the given time period.

| 5 to 10 years prior AD |  |  |  |  |
| --- | --- | --- | --- | --- |
|  | WOMEN |  | MEN |  |
|  | AGE(mean+/-sd) | N | AGE(mean+/-sd) | N |
| AD | 83.01+/-7.96 | 60251 | 80.62+/-8.14 | 34681 |
| CTL | 82.96+/-7.96 | 78100 | 80.55+/-8.15 | 42997 |

| 10 to 20 years prior AD |  |  |  |  |
| --- | --- | --- | --- | --- |
|  | WOMEN |  | MEN |  |
|  | AGE(mean+/-sd) | N | AGE(mean+/-sd) | N |
| AD | 82.99+/-7.98 | 52140 | 80.40+/-8.21 | 28535 |
| CTL | 82.94+/-7.99 | 64789 | 80.31+/-8.21 | 34292 |

### Associations

The associations between the biometric and blood borne measures and AD status were defined as Cohen’s effect sizes. For the prediction model, the data was split into 70:30 training and validation sets, and a random forest model based on 15 measures plus age and sex, selected based on a cohort coverage of at least half of those with blood pressure readings, was built and fit to the training data. The model was then tested on the validation set. The random forest analysis was carried out within the R environment using the randomForest package[26]. Predictability was assessed through a ROC AUC analysis.

The associations of drug prescriptions with AD incidence were defined by the corresponding odds ratios, where drug prescription status was a binary call based on any prescription of the drug over the given time period. For drugs mostly prescribed for one sex, based on prescription frequency being above 10-fold higher in men versus women or women versus men, analysis was restricted to the given sex. To test for the robustness of cohort matching we also calculated odds ratios with age, sex and number of observations as covariates with a linear logistic regression model. As a further sensitivity analysis, we included clinical practice as a random effect in the logistic regression model, we found that the inclusion of this error term and additional covariates had no significant effect on our conclusions.

## Results

### Biometric associations with AD incidence

Certain biometrics, like hypertension[27–29] and BMI[6], and blood borne marker levels[30–33] are associated with increased risk of developing AD with differences between people going on to develop AD and those that aren’t sometimes arising a relatively long time prior to diagnosis. As can be seen in Table 3 SBP is significantly high in the AD cohort 10 years prior to diagnosis, with an effect size of 0.89. Here, we find SBP in the AD cohort of 140.78+/-11.80 compared to 128.38+/-14.87 in the controls. Where we defined the AD cohort to mean those who will go on to develop the disease within the time frame of our dataset. The positive association of high SBP is consistent from ten years prior to diagnosis whereas the positive association of diastolic blood pressure (DBP) is more marked at ten years prior diagnosis than at later stages, with an effect size of 0.36 10 to 20 years prior index dropping to 0.05 in the 5 to 10 years range. Another previously reported association with AD incidence is blood serum albumin level[34]. This observation is validated in our dataset with albumin levels being the strongest negative association with AD incidence across the ten years prior to diagnosis, with levels of 41.97+/- 2.80 in the AD cohort and 42.82+/-3.04 in the controls. This association becomes stronger as diagnosis gets closer going from an effect of −0.27 at 10 to 20 years and −0.34 at 5 to 10 years prior to index.

**Table 3.** Most significant associations measured by Cohen’s d effect size with confidence intervals for blood measures and biometrics with AD for data averaged over 10 to 20 and 5 to 10 years prior to diagnosis.

| MEASURE | 10 to 20 years prior AD |  |  |  | 5 to 10 years prior AD |  |  |  |
| --- | --- | --- | --- | --- | --- | --- | --- | --- |
|  | Effect | Z | Controls | Cases | Effect | Z | Controls | Cases |
| Systolic arterial pressure | 0.89 (0.88 0.89) | 380.74 | 532574 | 206250 | 0.78 (0.78 0.79) | 377.44 | 702844 | 232436 |
| Diastolic arterial pressure | 0.36 (0.35 0.36) | 154.35 | 532574 | 206250 | 0.05 (0.04 0.05) | 22.82 | 702844 | 232436 |
| Serum urea level | 0.53 (0.52 0.54) | 146.27 | 199142 | 126440 | 0.62 (0.61 0.62) | 210.44 | 379119 | 185311 |
| Serum creatinine level | 0.37 (0.37 0.38) | 115.15 | 260245 | 159511 | 0.45 (0.45 0.46) | 160.94 | 431660 | 206344 |
| Serum albumin level | -0.27 (-0.28 -0.27) | -82.33 | 227793 | 141770 | -0.34 (-0.34 -0.33) | -124.69 | 400235 | 196314 |
| BMI | 0.18 (0.18 0.19) | 64.61 | 385426 | 158549 | 0.07 (0.07 0.08) | 26.21 | 458876 | 162233 |
| Plasma glucose level | 0.24 (0.23 0.25) | 52.34 | 123529 | 80656 | 0.25 (0.24 0.26) | 67.88 | 225115 | 120762 |
| Serum ferritin level | 0.37 (0.36 0.39) | 46.07 | 44579 | 25685 | 0.31 (0.30 0.32) | 61.58 | 121789 | 66178 |
| Serum testosterone level | 0.94 (0.90 0.99) | 45.77 | 10188 | 2709 | 0.92 (0.88 0.96) | 50.62 | 19029 | 3068 |
| Serum alkaline phosphatase level | 0.16 (0.15 0.16) | 45.42 | 222549 | 138334 | 0.17 (0.17 0.18) | 61.54 | 381972 | 188020 |
| Neutrophil count | -0.12 (-0.13 -0.12) | -40.68 | 275019 | 143771 | 0.02 (0.01 0.02) | 7.51 | 462694 | 202926 |
| Platelet count | -0.13 (-0.13 -0.12) | -39.57 | 287296 | 148334 | -0.15 (-0.15 -0.14) | -54.01 | 462701 | 202390 |
| Plasma urea level | 0.53 (0.51 0.56) | 38.38 | 12753 | 8976 | 0.55 (0.53 0.57) | 47.21 | 21237 | 12034 |
| Total white cell count | -0.12 (-0.12 -0.11) | -38.23 | 285247 | 147181 | -0.03 (-0.03 -0.02) | -11.4 | 464363 | 202847 |
| Serum glucose level | 0.31 (0.29 0.32) | 35.98 | 34341 | 24297 | 0.30 (0.29 0.32) | 42.33 | 56417 | 32344 |
| ALT/SGPT serum level | -0.12 (-0.13 -0.11) | -32.76 | 163690 | 109470 | -0.24 (-0.25 -0.24) | -76.04 | 236624 | 130858 |
| Plasma fasting glucose level | 0.18 (0.17 0.19) | 30.66 | 67846 | 51291 | 0.17 (0.16 0.18) | 40.53 | 153285 | 86210 |
| Plasma creatinine level | 0.40 (0.37 0.42) | 29.71 | 14514 | 9566 | 0.43 (0.41 0.45) | 40.46 | 26926 | 14196 |
| Monocyte count | 0.08 (0.08 0.09) | 26.55 | 273303 | 142968 | 0.20 (0.20 0.21) | 75.9 | 462225 | 202827 |
| Serum thyroid stimulating hormone level | 0.10 (0.09 0.11) | 24.96 | 183629 | 105520 | 0.09 (0.08 0.09) | 29.35 | 367896 | 172837 |

Given that there are multiple factors showing association with AD incidence we sought to build a predictive model based on these factors. As can be seen in Table 3, there is variable coverage of the cohort across the measures. Restricting measures defined in the 10 to 20 years prior to index with cohort coverage above half of the SBP we are left with 15 measures defined for a cohort of 219,639.

Splitting this data into 70:30 training and validation sets and training a random forest predictor model on the training set we find a ROC AUC (mean+/-sd) of 0.79+/-0.001 from 10 independent data split runs (accuracy 0.71+/-0.001, precision 0.78+/-0.002, recall 0.66+/-0.003). Not surprisingly, this modest level of predictability falls far short of models based on AD specific biomarkers such as phosphorylated tau and amyloid[35]. However, in the next section we will see how the observational correlates of AD incidence can help in the interpretation of epidemiological drug repurposing.

### Medication prescription associations with AD incidence

As described in the Methods section, the study cohort was filtered based on matching prescription and observation frequencies over given time frames prior to index date. To see to what extent prior drug use is associated with AD incidence we gathered prescription data in time frames of 5 to 10 and 10 to 20 years prior to diagnosis. For sex specific drugs analysis was restricted to the given sex. The top positive and negative associations of drug prescriptions prior to index with AD incidence are shown in Table 4.

**Table 4.** The associations of prior drug prescriptions with AD incidence. The top 20 positive and negative associations are shown (ranked by Z score) for drugs taken by both sexes.

| DRUG | 5-10 years prior AD |  | 10-20 years prior AD |  | DRUG | 5-10 years prior AD |  | 10-20 years prior AD |  |
| --- | --- | --- | --- | --- | --- | --- | --- | --- | --- |
|  | logODDs | Z | logODDs | Z |  | logODDs | Z | logODDs | Z |
| PHENOXYMETHYLPENICILLIN | -0.90 (-0.93 -0.87) | -52.02 | -0.93 (-0.96 -0.90) | -59.59 | ASPIRIN | 1.44 (1.42 1.46) | 124.98 | 0.90 (0.88 0.93) | 67.96 |
| CLINDAMYCIN | -1.48 (-1.57 -1.38) | -31.07 | -1.65 (-1.76 -1.54) | -29.02 | SIMVASTATIN | 1.24 (1.22 1.26) | 115.11 | 0.75 (0.72 0.77) | 53.92 |
| SUMATRIPTAN | -1.28 (-1.38 -1.17) | -24.43 | -0.89 (-1.00 -0.79) | -17.06 | PARACETAMOL | 0.88 (0.86 0.90) | 92.15 | 0.31 (0.29 0.33) | 30.23 |
| LYMECYCLINE | -1.28 (-1.39 -1.18) | -23.18 | -1.27 (-1.43 -1.11) | -15.46 | BENDROFLUMETHIAZIDE | 1.17 (1.14 1.19) | 90.62 | 0.73 (0.71 0.76) | 56.13 |
| FLUOXETINE | -0.50 (-0.54 -0.46) | -22.8 | -0.65 (-0.69 -0.61) | -30.46 | AMLODIPINE | 1.05 (1.02 1.07) | 78.52 | 0.69 (0.65 0.72) | 38.38 |
| MEBENDAZOLE | -1.68 (-1.84 -1.53) | -21.18 | -1.80 (-1.94 -1.67) | -25.64 | CALCIUM CARBONATE | 1.08 (1.05 1.11) | 77.69 | 0.68 (0.64 0.72) | 34.74 |
| ORLISTAT | -0.89 (-0.97 -0.81) | -20.9 | -0.92 (-1.01 -0.83) | -19.86 | FUROSEMIDE | 1.33 (1.29 1.36) | 76.05 | 0.65 (0.61 0.69) | 32.22 |
| VARENICLINE | -0.93 (-1.02 -0.84) | -20.03 | -0.96 (-1.11 -0.81) | -12.39 | COLECALCIFEROL | 1.03 (1.01 1.06) | 73.03 | 0.79 (0.74 0.83) | 34.85 |
| FLUCONAZOLE | -0.50 (-0.55 -0.46) | -19.91 | -0.73 (-0.78 -0.68) | -26.12 | ATORVASTATIN | 1.00 (0.97 1.03) | 68.62 | 0.64 (0.61 0.68) | 33.61 |
| NICOTINE | -0.50 (-0.55 -0.44) | -18.94 | -0.53 (-0.59 -0.48) | -19.79 | RAMIPRIL | 0.91 (0.89 0.94) | 67.08 | 0.52 (0.49 0.56) | 28.18 |
| CYPROTERONE | -1.59 (-1.77 -1.41) | -17.5 | -2.69 (-2.92 -2.47) | -23.08 | ATENOLOL | 1.05 (1.02 1.08) | 65.8 | 0.62 (0.59 0.65) | 42.21 |
| PRILOCAINE | -1.51 (-1.69 -1.35) | -17.22 | -1.62 (-1.83 -1.43) | -15.84 | HYPROMELLOSE | 1.32 (1.28 1.36) | 65.1 | 0.97 (0.92 1.02) | 42.1 |
| ERYTHROMYCIN | -0.24 (-0.27 -0.21) | -16.99 | -0.43 (-0.45 -0.40) | -33.46 | CO CODAMOL | 0.62 (0.60 0.64) | 61.71 | 0.31 (0.29 0.33) | 27.36 |
| PROGUANIL | -0.68 (-0.77 -0.60) | -16.52 | -0.29 (-0.37 -0.22) | -7.32 | BISOPROLOL | 1.06 (1.03 1.10) | 57.02 | 0.65 (0.59 0.71) | 21.17 |
| ATOVAQUONE | -0.68 (-0.77 -0.60) | -16.29 | -0.29 (-0.38 -0.21) | -6.83 | OMEPRAZOLE | 0.59 (0.57 0.61) | 56.87 | 0.34 (0.31 0.37) | 24.61 |
| SALBUTAMOL | -0.17 (-0.20 -0.15) | -15.28 | -0.50 (-0.52 -0.47) | -37.43 | PARAFFIN | 0.60 (0.58 0.62) | 56.65 | 0.14 (0.11 0.16) | 10.99 |
| ADAPALENE | -3.50 (-4.02 -3.05) | -14.3 | -3.27 (-3.93 -2.72) | -10.69 | MACROGOL | 0.81 (0.78 0.84) | 54.43 | 0.33 (0.28 0.38) | 12.97 |
| ALUMINIUM CHLORIDE | -1.49 (-1.71 -1.27) | -13.28 | -1.71 (-1.97 -1.46) | -12.95 | QUININE | 1.25 (1.20 1.29) | 53.66 | 0.90 (0.85 0.95) | 35.18 |
| PROPRANOLOL | -0.32 (-0.37 -0.27) | -13.18 | -0.27 (-0.32 -0.23) | -11.8 | WARFARIN | 1.27 (1.22 1.32) | 51.91 | 0.63 (0.56 0.69) | 19.61 |
| CLOTRIMAZOLE | -0.17 (-0.19 -0.14) | -12.9 | -0.41 (-0.44 -0.38) | -29.8 | LACTULOSE | 0.78 (0.75 0.81) | 50.93 | 0.21 (0.18 0.25) | 12.48 |

In agreement with our observations on the AD risk associated with high blood pressure, we find enhanced prescriptions for anti-hypertensive medications (bendroflumethiazide, amlodipine, furosemide, atenolol, bisoprolol, ramipril) in the AD cohort, see Table 4 on the right. Of interest in the negative association drug set is Salbutamol which has previously been reported to protect against PD based on epidemiological evidence[17], but this has been disputed based on confounding effects of smoking and the protective effects of nicotine[36]. Interestingly, salbutamol has been hypothesised as a PD repurposing candidate based its driving the transcription of the FGF20[13]. Smoking has been reported to be associated with increased rates of dementia[37, 38], ranking number three in the set of modifiable risks[39]. However, there have been reports on the possible cognitive benefits of nicotine[40, 41]. In this context, the present analysis shows a negative association of nicotine with AD incidence, and it would be interesting to include data on non-prescription nicotine use to see to what extent the effects of salbutamol and nicotine can be separated.

### Type 2 diabetes drugs

Not shown in Table 4 are the GLP-1 agonist and SGLT2 inhibitor class of T2D medications which have consistent negative associations with AD incidence, see Table 7 for details. Given the consistent negative association we also defined correlations for prescription of any of the GLP-1 drugs in the data base, see entries under pooled in Table 7. Interestingly, the GLP-1 agonist liraglutide has been recently reported to have positive effects in a year-long phase 2 trial in people with AD[42] and the closely related exenatide has shown evidence of reducing AD risk[43]. Other groups have reported on the positive effects of GLP-1 agonists in PD. This result is of interest outside diabetes as a GLP-1 medication, semaglutide, has recently been licensed as a weight loss drug[44, 45]. It will be of great interest to follow dementia incidence in this population. Another class of T2D medication showing potentially positive effects are the glucose lowering SGLT2 inhibitors, see Table 7. This has also been reported in a recent systematic review[46]. The SGLT2 inhibitors have only been licenced relatively recently and we thus did not have data to power an analysis 10 years prior to diagnosis of AD. These results are of especial interest as T2D is a known risk factor for developing the disease[22, 47–49]. In agreement with this, our data show a positive association with the most frequent T2D medication, metformin, and AD incidence, (logODDs 0.21 (0.16 0.27) Z 7.9 at 10 to 20 years prior AD incidence and logODDs 0.69 (0.65 0.73) Z 37.67at 5 to 10 prior AD incidence). The observational data enables us to uncouple the effects of disease and the drugs for which they are prescribed. To this end we delimited T2D disease cohorts based on a T2D diagnosis prior to AD onset. In the 10 to 20 years prior AD diagnosis medication dataset there are 6,822 men aged 80.58+/-7.78 and 12,567 women aged 82.68+/-7.70 with a T2D observation as described in Methods. In the 5 to 10 years prior AD diagnosis medication dataset there are 8,415 men aged 80.62+/-7.69 and 14,505 women aged 82.53+/-7.73 with a T2D observation as described in Methods. In agreement with T2D being a risk factor for AD, we find a higher future AD incidence in those with a T2D observation, within these time frames. In particular, 10 to 20 years prior to AD diagnosis 48% of those with T2D will go on to develop AD as opposed to 45% in those with no T2D observation, with an effect size of 0.14 (0.11 0.17) Z 9.02. Whereas 5 to 10 years prior AD onset there is a larger risk associated with T2D, with 54% of those with T2D going on to develop AD and only 43% in the population without an T2D diagnosis. This observation bolsters the positive effects of the T2D drugs as they show a negative association with AD and are prescribed for a condition that is associated with a higher AD risk. Further, restricting the analysis to the T2D cohort we find a stronger negative association with AD incidence with the GLP-1 agonists and SGLT2 antagonist drugs, see Table 7.

### Triptans and migraine

Various triptan migraine medications appear to negatively associate with AD incidence. The correlations for the complete set of triptan drugs are shown in Table 7 together with the correlations for a prescription of any of the triptans. In contrast to T2D discussed above, there is no consensus in the literature as to whether migraine is a risk factor for AD, with groups reporting contradictory results [50, 51]. Interestingly, there may be an association of blood pressure with migraine incidence, with a large study showing a positive association of low blood pressure and migraine incidence in adolescents[52]. However, there does not appear to be a consensus on whether migraine is due to high or low blood pressure or whether there is indeed any association[53]. In this context, our data shows that those on the triptan class of migraine medication have significantly lower SBP. For, example at 10 to 20 years prior to AD diagnosis SBP is lower in the triptan cohort in both the control and case cohorts, with effect sizes of −0.25 (−0.29 −0.21) N triptans +/- 2,477/ 68,855 in the controls and −0.23 (−0.30 −0.16) N triptans +/- 834/ 65655 in cases. Similarly, at 5 to 10 years prior to AD diagnosis SBP is lower in the triptan cohort in both the control and case cohorts, with effect sizes of −0.29 (−0.33 −0.25) N triptans +/- 3,067/ 91,171 in the controls and −0.17 (−0.24 −0.09) N triptans +/- 727/87,702 in cases. Triptans make up only for part of the migraine prescription options, with NSAIDs being another option, see the BNF drug data (www-medicinescomplete-com.apollo.worc.ac.uk/#/browse/bnf). The observational data allows for delimiting a migraine cohort within the AD case control cohort. The overall migraine incidence 10 to 20 years prior to index is 3.5%. However, we find that this rate is significantly lower, 2.07%, in the those going on to develop AD, with a log odds of −0.73 (−0.76 −0.69) and Z score of −38.62. Restricting to the migraine cohort, we observe a much diminished effect size of the triptan medication association with AD incidence, both at 5 to 10 and 10 to 20 years prior diagnosis, see Table 7. In particular, we observe that 50% and 72% of the strength of triptan association with AD incidence is explained by migraine diagnosis at 5 to 10 and 10 to 20 years prior AD diagnosis respectively. This is in contrast to the anti-diabetic medications where the negative association with AD incidence was strengthened when restricting analysis to the T2D cohort. A properly matched migraine case/control study would have to be performed to see whether the residual negative association of triptan medication with AD incidence still obtains. This also goes for establishing a rigorous association between SBP and AD incidence.

### Hormonal drugs

In the analysis of drugs predominantly prescribed to women, see Table 5, we find consistent negative associations with hormonal medications with the top negative association with AD incidence at both 5 to 10 years and 10 to 20 years prior index being with the postmenopausal medication estradiol. In this context the protective effects of oestrogen in the context of AD it has been reported that postmenopausal women on oestrogen had significantly delayed AD onset[54, 55]. However, a trial in AD patients did not show a positive effect of disease progression, rather the reverse[56, 57]. It has been suggested that the positive effects of hormone replacement are only present in a younger cohort[58]. Prescription of the weaker estrogen estriol is associated with increased AD incidence, and this may be accounted for by the types of conditions it is prescribed for, such as urinary tract infections and vaginal atrophy in contrast with the hormonal replacement use of the negatively associated estrogen drugs.

**Table 5.** The associations of prior drug prescriptions with AD incidence. The top 20 positive and negative associations are shown (ranked by Z score) for drugs taken predominantly by women.

| DRUG | 5-10 years prior AD |  | 10-20 years prior AD |  | DRUG | 5-10 years prior AD |  | 10-20 years prior AD |  |
| --- | --- | --- | --- | --- | --- | --- | --- | --- | --- |
|  | logODDs | Z | logODDs | Z |  | logODDs | Z | logODDs | Z |
| ESTRADIOL | -1.57 (-1.61 -1.53) | -77.47 | -1.26 (-1.30 -1.23) | -73.57 | ESTRIOL | 0.66 (0.60 0.72) | 21.03 | 0.66 (0.59 | 20.09 |
| NORETHISTERONE | -2.43 (-2.51 -2.34) | -55.94 | -1.45 (-1.50 -1.40) | -60.07 | STRONTIUM | 1.57 (1.39 1.76) | 16.89 | 0.85 (0.58 | 6.12 |
| PROGESTERONE | -2.05 (-2.16 -1.94) | -35.46 | -0.98 (-1.05 -0.92) | -30.13 | IBANDRONIC | 1.10 (0.94 1.26) | 13.33 | 0.91 (0.65 | 6.62 |
| LEVONORGESTREL | -5.33 (-5.64 -5.05) | -35.33 | -4.16 (-4.32 -4.01) | -52.55 | ANASTROZOLE | 0.62 (0.49 0.75) | 9.42 | 0.41 (0.21 | 4.12 |
| MEDROXYPROGESTERONE | -2.02 (-2.13 -1.90) | -34.72 | -0.95 (-1.01 -0.88) | -28.58 | TAMOXIFEN | 0.51 (0.40 0.62) | 9 | 0.61 (0.51 | 11.92 |
| MEFENAMIC | -2.93 (-3.11 -2.75) | -32.02 | -2.15 (-2.26 -2.05) | -40.5 | RALOXIFENE | 0.99 (0.76 1.24) | 8.12 | 0.74 (0.54 | 7.43 |
| ETHINYLESTRADIOL | -6.18 (-6.62 -5.80) | -29.63 | -4.44 (-4.61 -4.28) | -53.79 | LETROZOLE | 0.63 (0.46 0.80) | 7.36 | 0.48 (0.11 | 2.55 |
| TRANEXAMIC | -3.06 (-3.27 -2.86) | -28.95 | -2.31 (-2.45 -2.18) | -33.15 | DENOSUMAB | 1.34 (0.93 1.77) | 6.32 | 0.22 (-3.01 | 0.15 |
| DESOGESTREL | -5.54 (-6.00 -5.14) | -25.34 | -4.22 (-4.52 -3.94) | -28.44 | EXEMESTANE | 0.47 (0.16 0.79) | 2.97 | 0.29 (-0.24 | 1.07 |
| ETONOGESTREL | -5.29 (-6.32 -4.52) | -11.83 | -4.28 (-5.31 -3.51) | -9.54 | ALENDRONIC | 1.73 (0.59 3.20) | 2.7 | NA | NA |
| EFLORNITHINE | -1.17 (-1.49 -0.88) | -7.52 | -0.70 (-1.11 -0.31) | -3.45 | TERIPARATIDE | 1.65 (0.26 3.54) | 2.08 | NA | NA |
| CO.CYPRINDIOL | -5.83 (-8.69 -4.34) | -5.91 | -5.16 (-8.02 -3.67) | -5.15 |  |  |  |  |  |
| NONOXINOL | -2.18 (-3.13 -1.43) | -5.13 | -2.35 (-2.81 -1.94) | -10.62 |  |  |  |  |  |
| TIBOLONE | -0.34 (-0.48 -0.20) | -4.62 | 0.20 (0.11 0.28) | 4.52 |  |  |  |  |  |
| PAPILLOMAVIRUS | -4.02 (-6.89 -2.51) | -3.99 | -3.45 (-6.32 -1.92) | -3.41 |  |  |  |  |  |
| OESTROGENS | -0.11 (-0.19 -0.02) | -2.5 | 0.21 (0.16 0.26) | 7.77 |  |  |  |  |  |

The number of male specific drugs is smaller than those prescribed specifically for women, and we find no significant negative associations with AD incidence and prescription of these medications. The drugs associated with increased AD incidence are prescribed for prostate enlargement and impotence, see Table 6. It is not clear how these medications of the corresponding diseases are linked to AD, though it has been reported that men with benign prostate hyperplasia have a higher rate of AD and other dementias[59].

**Table 6.** The associations of prior drug prescriptions with AD incidence. The top 20 positive and negative associations are shown (ranked by Z score) for drugs taken predominantly by men.

| DRUG | 5-10 years prior AD |  | 10-20 years prior AD |  |
| --- | --- | --- | --- | --- |
|  | logODDs | Z | logODDs | Z |
| TAMSULOSIN | 1.35 (1.30 1.41) | 47.55 | 0.95 (0.87 1.03) | 23.26 |
| FINASTERIDE | 1.43 (1.34 1.51) | 32.31 | 0.98 (0.86 1.11) | 15.67 |
| ALFUZOSIN | 1.25 (1.12 1.38) | 18.44 | 0.79 (0.64 0.94) | 10.33 |
| SILDENAFIL | 0.48 (0.42 0.54) | 15.95 | 0.47 (0.40 0.53) | 13.43 |
| DUTASTERIDE | 1.33 (1.17 1.51) | 15.51 | 0.95 (0.69 1.23) | 7 |
| BICALUTAMIDE | 1.44 (1.23 1.67) | 12.89 | 0.89 (0.58 1.21) | 5.48 |
| TADALAFIL | 0.51 (0.43 0.60) | 12.32 | 0.48 (0.37 0.59) | 8.54 |
| INDORAMIN | 1.49 (1.21 1.79) | 10.04 | 0.93 (0.74 1.14) | 9.14 |
| VARDENAFIL | 0.62 (0.46 0.77) | 7.82 | 0.33 (0.14 0.52) | 3.46 |
| APOMORPHINE | 1.50 (0.95 2.10) | 5.13 | 0.53 (0.25 0.82) | 3.63 |
| TRIPTORELIN | 1.41 (0.89 1.98) | 5.09 | NA | NA |
| ALPROSTADIL | 0.47 (0.21 0.74) | 3.47 | 0.70 (0.44 0.96) | 5.27 |

**Table 7.** Sets of compounds belonging to classes. The associations based on prescriptions of any of the medications in the various classes are shown as combined scores.

| TRIPTANS |  |  |  |  | SGLT2 antagonists |  |  |  |  |
| --- | --- | --- | --- | --- | --- | --- | --- | --- | --- |
|  | 5 to 10 years |  | 10 to 20 years |  |  | 5 to 10 years |  | 10 to 20 years |  |
|  | logODDS | Z | logODDS | Z |  | logODDS | Z | logODDS | Z |
| ALMOTRIPTAN | -1.03 (-1.41 -0.67) | -5.44 | -1.15 (-1.55 -0.77) | -5.8 | CANAGLIFLOZIN | -0.34 (-1.00 0.27) | -1.07 | NA | NA |
| ELETRIPTAN | -2.32 (-5.22 -0.71) | -2.24 | -0.89 (-2.40 0.32) | -1.34 | DAPAGLIFLOZIN | -0.86 (-1.26 -0.48) | -4.32 | NA | NA |
| FROVATRIPTAN | -1.61 (-2.20 -1.10) | -5.82 | -1.06 (-1.62 -0.56) | -3.99 | EMPAGLIFLOZIN | -1.48 (-2.56 -0.61) | -3.05 | NA | NA |
| NARATRIPTAN | -1.22 (-1.48 -0.98) | -9.67 | -0.95 (-1.14 -0.78) | -10.33 | **POOLED** | -0.83 (-1.16 -0.52) | -5.12 | NA | NA |
| RIZATRIPTAN | -1.17 (-1.37 -0.98) | -12 | -0.97 (-1.16 -0.79) | -10.28 | **T2D cohort** | -1.16 (-1.51 -0.83) | -6.65 | NA | NA |
| SUMATRIPTAN | -1.28 (-1.38 -1.17) | -24.43 | -0.89 (-1.00 -0.79) | -17.06 |  |  |  |  |  |
| ZOLMITRIPTAN | -1.40 (-1.62 -1.19) | -12.67 | -0.90 (-1.08 -0.73) | -10.36 |  |  |  |  |  |
| **POOLED** | -1.24 (-1.32 -1.16) | -29.99 | -0.92 (-1.00 -0.84) | -23.04 |  |  |  |  |  |
| **MIGRAINERS** | -0.62 (-0.76 -0.49) | -8.88 | -0.26 (-0.38 -0.14) | -4.29 |  |  |  |  |  |
| GLP1 agonists |  |  |  |  | Hormonals |  |  |  |  |
|  | 5 to 10 years |  | 10 to 20 years |  |  | 5 to 10 years |  | 10 to 20 years |  |
|  | logODDS | Z | logODDS | Z |  | logODDS | Z | logODDS | Z |
| DULAGLUTIDE | -0.60 (-2.14 0.68) | -0.88 | NA | NA | DESOGESTREL | -5.54 (-6.00 -5.14) | -25.34 | -4.22 (-4.52 -3.94) | -28.44 |
| EXENATIDE | -0.47 (-0.78 -0.16) | -2.96 | -0.96 (-1.77 -0.26) | -2.53 | ESTRADIOL | -1.57 (-1.61 -1.53) | -77.47 | -1.26 (-1.30 -1.23) | -73.57 |
| LIRAGLUTIDE | -0.45 (-0.74 -0.17) | -3.07 | -1.02 (-2.13 -0.09) | -2 | ETHINYLESTRADIOL | -6.18 (-6.62 -5.80) | -29.63 | -4.44 (-4.61 -4.28) | -53.79 |
| **POOLED** | -0.41 (-0.63 -0.20) | -3.73 | -0.97 (-1.66 -0.36) | -2.95 | ETONOGESTREL | -5.29 (-6.32 -4.52) | -11.83 | -4.28 (-5.31 -3.51) | -9.54 |
| **T2D cohort** | -0.75 (-0.98 -0.52) | -6.26 | -1.45 (-2.29 -0.74) | -3.72 | LEVONORGESTREL | -5.33 (-5.64 -5.05) | -35.33 | -4.16 (-4.32 -4.01) | -52.55 |
|  |  |  |  |  | MEDROXYPROGESTERONE | -2.02 (-2.13 -1.90) | -34.72 | -0.95 (-1.01 -0.88) | -28.58 |
|  |  |  |  |  | NORETHISTERONE | -2.43 (-2.51 -2.34) | -55.94 | -1.45 (-1.50 -1.40) | -60.07 |
|  |  |  |  |  | OESTROGENS | -0.11 (-0.19 -0.02) | -2.5 | 0.21 (0.16 0.26) | 7.77 |
|  |  |  |  |  | PROGESTERONE | -2.05 (-2.16 -1.94) | -35.46 | -0.98 (-1.05 -0.92) | -30.13 |
|  |  |  |  |  | **POOLED** | -1.94 (-1.97 -1.90) | -107.8 | -1.36 (-1.38 -1.33) | -91.39 |

## Conclusions

Effective disease modifying therapeutics for AD have so far proved elusive due to the multi-factorial nature of the condition with a complex set of diverse causes converging on catastrophic dementia with onerous and increasing burdens on the health care system as well as the obvious toll it takes on the quality of life for the individual. The increasing availability of curated diagnosis, prescription and biometric data, through resources such as the CPRD[19], HDRUK[18] and UK Biobank[25], has allowed for an investigation of the associations between modifiable lifestyle factors and disease incidence that can inform guidelines as well as providing a basis for segregating populations for focussed early intervention. Also, strong associations between drug use and disease incidence can build hypotheses for repurposing initiatives. The present study sought to investigate both biometric and prescription associations with future AD incidence within one case-control cohort, where each case is demographically matched with a control at the time of diagnosis and data collected prior to diagnosis.

Our findings recapitulate several significant associations between biometric measures and the risk of developing AD. Specifically, SBP and DBP were consistently associated with an increased risk of AD, with SBP showing a significant positive association up to ten years before diagnosis. Blood serum albumin levels also emerged as a strong negative predictor of AD, reaffirming its potential role as a biomarker for early detection. It would be of interest to see whether compromised blood brain barrier due to elevated blood pressure[60–62] and reduced clearance of toxic protein aggregates due to low albumin levels[63–65] contribute additively to dementia risk. There may be however and independent correlation between serum albumin and hypertension, with a study showing that low albumin levels predict hypertension[66]. Supporting this, out of all the blood borne markers we have data for, serum albumin level is the most significant negative correlator of SBP level (logODDs −0.20 Z −37.93, controlling for age and sex). This motivates further investigation of dementia incidence in people with hypoalbuminemia.

In terms of medication prescriptions, our analysis dovetails with the biometric data, showing a relatively high prescription of anti-hypertensives in the cohort going on to develop AD. As such, the drug associations reflect the risk associated with hypertension and not with taking the drug. Of the drugs showing a negative association with AD, salbutamol has been shown to be protective in PD in a relatively large epidemiological study[17] and an animal model of the disease[13], but the direct causal link has been questioned with a study claiming that the association is confounded by the relatively low PD incidence in smokers[36]. In contrast, there appears to be a consensus that smoking is a dementia risk[39], and it is difficult to argue that smoking would confound the negative association of salbutamol use with AD incidence. However, we see a negative association of nicotine prescription and AD incidence and it would be of interest to include non-prescription nicotine use in the analysis to see whether the salbutamol effect is robust. Other drugs with reported beneficial effects in dementia are two classes of T2D medications, the insulin modulating GLP-1 agonists and the glucose lowering SGLT2 inhibitors. These results are particularly interesting given the increased dementia incidence in diabetics[67]. The SGLT2 inhibitors have only been licenced relatively recently and we thus did not have data to power an analysis 10 years prior to diagnosis of AD and it will be of interest to return to the analysis once more long-term use data becomes available. The GLP-1 agonists have a longer track record as T2D medications but perhaps of greater interest is the recent licencing of the GLP-1 agonist semaglutide as a weight loss drug[68]. The subsequent larger cohort of users will facilitate a more robust association analysis. The strongest negative associations with AD incidence in AD were with postmenopausal hormonal prescriptions. The protective effects of estrogen has been a long established[54, 55], however the subsequent clinical trials proved to be disappointing[56, 57]. One criticism of the trial is that it was conducted with older women and perhaps therefore less relevant to investigating a protective effect[58]. Apart from the reported neuroprotective effects of sumatriptan in an animal stroke model[69], as far as we know the triptan class of migraine medications have not previously been reported in connection with neurodegenerative conditions. Our analysis points to the negative association of triptan prescription with AD incidence as being condition driven, as we observe a lower AD incidence in the migraine population. Further, when the analysis is restricted to those with a migraine diagnosis this association is significantly reduced. This observation highlights the importance of the inclusion of concurrent observational data in drug repurposing strategies based on prescription data.

Overall, these results underscore the importance of a multifaceted approach in understanding and addressing intervention in AD. By integrating observational data and medication histories, we can enhance our ability to predict and potentially mitigate AD risk through tailored interventions. Future research should continue to explore these associations in larger and more diverse populations, considering the potential for drug repurposing and the development of targeted preventive strategies.

### Study limitations

As with most retrospective studies, it is problematic to make conclusions as to causality based on correlations. For example, as shown here, the seemingly protective effects of anti-migraine medication are confounded by the lower incidence of AD in the migraine cohort. Which in turn may be due to the differential biometrics in the migraine cohort. In contrast to the analysis of biometrics and blood borne factors, where one is comparing numerical measures, the analysis of medication incidence is complicated by being categorical, where a null assignment for a given medication may be due to the lack of data. The present study aimed to circumvent this problem by filtering the cohort to ensure a degree of observation and prescription matching.

## Data Availability

The Clinical Practice Research Datalink data is available upon submission of a protocol

https://www.cprd.com/

## Acknowledgements

The author would like to thank Dr Alexadru Dregan for extracting the data from CPRD and helping with technical issues, and Professor Patrick Doherty for reading the paper and providing helpful suggestions. This study is based in part on data from the Clinical Practice Research Datalink obtained under licence from the UK Medicines and Healthcare products Regulatory Agency. The data is provided by patients and collected by the NHS as part of their care and support. The interpretation and conclusions contained in this study are those of the author alone.

## Ethical approval

The CPRD group obtained ethical approval from a National Research Ethics Service Committee for all purely observational research using anonymised CPRD data, as in the present study. The study protocol has been approved by the Independent Scientific Advisory Committee for Medicines and Healthcare Products Regulatory Agency, Ref: 14_111.

## Supplementary Data

**Table S1.** The SNOMED codes and descriptions for AD assignment in the case cohort. First ever diagnosis of AD was between 1/1/2012 and 31/12/2022 and this was defined as the index date for control cohort matching.

| Original Read Code | Term | Snomed CT Concept Id | Snomed CT Description Id |
| --- | --- | --- | --- |
| F110 | Alzheimer's disease | 26929004 | 45046017 |
| Eu00 | [X]Dementia in Alzheimer's disease | 26929004 | 45046017 |
| Eu002 | [X]Dementia in Alzheimer's dis, atypical or mixed type | 26929004 | 45046017 |
| Eu00z | [X]Dementia in Alzheimer's disease, unspecified | 26929004 | 45046017 |
| Fyu30 | [X]Other Alzheimer's disease | 26929004 | 45046017 |
| Eu00z-1 | [X]Alzheimer's dementia unspec | 26929004 | 45046017 |
| ^ESCTAD293123 | AD - Alzheimer's disease | 26929004 | 1225144019 |
| ^ESCTAL293124 | Alzheimer disease | 26929004 | 2839267017 |
| ^ESCTAL293125 | Alzheimer dementia | 26929004 | 3424952012 |
| Eu025 | Diffuse Lewy body disease | 80098002 | 132893017 |
| F116 | Lewy body disease | 80098002 | 132893017 |
| ^ESCTLE380262 | Lewy body variant of Alzheimer's disease | 80098002 | 132894011 |
| ^ESCTLB380264 | LBD - Lewy body disease | 80098002 | 1234436013 |
| ^ESCTDE380265 | Dementia of the Lewy body type | 80098002 | 1234437016 |
| ^ESCTCO380267 | Cortical Lewy body disease | 80098002 | 1234439018 |
| ^ESCTFA500550 | Familial Alzheimer's disease of early onset | 230265002 | 345083019 |
| ^ESCTFA500554 | Familial Alzheimer's disease of late onset | 230267005 | 345085014 |
| ^ESCTFO500558 | Focal Alzheimer's disease | 230269008 | 345087018 |
| F1100 | Alzheimer's disease with early onset | 416780008 | 2957141019 |
| Eu000-3 | [X]Alzheimer's disease type 2 | 416780008 | 2957119019 |
| Eu000 | Dementia in Alzheimer's disease with early onset | 416780008 | 2957119019 |
| Eu000-1 | [X]Presenile dementia,Alzheimer's type | 416780008 | 2957138011 |
| Eu000-2 | [X]Primary degen dementia, Alzheimer's type, presenile onset | 416780008 | 2957119019 |
| ^ESCTPR689721 | Primary degenerative dementia of the Alzheimer type, presenile onset | 416780008 | 2549310013 |
| ^ESCTDE689724 | Dementia of the Alzheimers type with early onset | 416780008 | 2957133019 |
| ^ESCTPR689725 | Presenile dementia, Alzheimer's type | 416780008 | 2957138011 |
| ^ESCTDE689727 | Dementia in Alzheimer's disease - type 2 | 416780008 | 2957156011 |
| F1101 | Alzheimer's disease with late onset | 416975007 | 2957124016 |
| Eu001-1 | [X]Alzheimer's disease type 1 | 416975007 | 2957137018 |
| Eu001 | Dementia in Alzheimer's disease with late onset | 416975007 | 2957137018 |
| Eu001-3 | [X]Primary degen dementia of Alzheimer's type, senile onset | 416975007 | 2957137018 |
| Eu001-2 | [X]Senile dementia,Alzheimer's type | 416975007 | 2957124016 |
| ^ESCTPR690018 | Primary degenerative dementia of the Alzheimer type, senile onset | 416975007 | 2549518019 |
| ^ESCTDE690020 | Dementia of the Alzheimers type, late onset | 416975007 | 2957123010 |
| ^ESCTSD690022 | SDAT - Senile dementia, Alzheimer's type | 416975007 | 2957134013 |
| ^ESCTDE690024 | Dementia in Alzheimer's disease - type 1 | 416975007 | 2957155010 |
| ^ESCTNO783407 | Non-amnestic Alzheimer disease | 722600006 | 3332846016 |
| ^ESCTMI802420 | Mixed dementia | 79341000119107 | 2955467016 |
| ^ESCT1237064 | Dementia with mixed etiology | 79341000119107 | 3670850017 |
| ^ESCT1237065 | MVAD - Mixed vascular Alzheimer dementia | 79341000119107 | 3670851018 |
| ^ESCT1237066 | Dementia with mixed aetiology | 79341000119107 | 3670852013 |
| ^ESCTDE804491 | Delusions in Alzheimer's disease | 141991000119109 | 2968442016 |
| ^ESCTDE804494 | Depressed mood in Alzheimer's disease | 142001000119106 | 2968437012 |
| ^ESCTNO500556 | Non-familial Alzheimer's disease of late onset | 230268000 | 345086010 |
| ^ESCTNO500552 | Non-familial Alzheimer's disease of early onset | 230266001 | 345084013 |
| ^ESCTAL804498 | Alzheimer's disease co-occurrent with delirium | 142011000119109 | 3311926019 |
| ^ESCTPR257573 | Primary degenerative dementia of the Alzheimer type, senile onset, with delirium | 4817008 | 9023015 |
| ^ESCTPR292993 | Primary degenerative dementia of the Alzheimer type, senile onset, with depression | 26852004 | 44934012 |
| ^ESCTEA803291 | Early onset Alzheimer's disease with behavioural disturbance | 105421000119105 | 2985868010 |
| ^ESCTEA803293 | Early onset Alzheimers disease with behavioral disturbance | 105421000119105 | 2987421013 |
| ^ESCTDE796928 | Dementia of the Alzheimer type with behavioural disturbance | 1581000119101 | 2967544019 |
| ^ESCTPR357504 | Primary degenerative dementia of the Alzheimer type, senile onset, uncomplicated | 66108005 | 109791011 |
| ^ESCTPR260327 | Primary degenerative dementia of the Alzheimer type, presenile onset, uncomplicated | 6475002 | 11739017 |
| EMISNQDD3 | [D] Dementia in Alzheimer's disease | 914951000006105 | 914951000006114 |
| EMISICD10\|F0024 | Dementia in Alzheimer's dis, atypical or mixed type, other mixed symptoms | 1972341000006107 | 1972341000006111 |
| EMISICD10\|F0094 | Dementia in Alzheimer's disease, unspecified, other mixed symptoms | 1972471000006107 | 1972471000006111 |
| EMISICD10\|F0020 | Dementia in Alzheimer's dis, atypical or mixed type, without additional symptoms | 1972231000006105 | 1972231000006114 |
| HNG0062 | [RFC] Alzheimer's disease | 905791000006104 | 905791000006115 |
| EMISICD10\|F0014 | Dementia in Alzheimer's disease with late onset, other mixed symptoms | 1972211000006104 | 1972211000006115 |
| EMISICD10\|F0004 | Dementia in Alzheimer's disease with early onset, other mixed symptoms | 1972141000006109 | 1972141000006113 |
| EMISICD10\|F0090 | Dementia in Alzheimer's disease, unspecified, without additional symptoms | 1972371000006104 | 1972371000006115 |
| EMISICD10\|F0010 | Dementia in Alzheimer's disease with late onset, without additional symptoms | 1972171000006101 | 1972171000006117 |
| EMISICD10\|F0022 | Dementia in Alzheimer's dis, atypical or mixed type, other symptoms, predominantly hallucinatory | 1972291000006109 | 1972291000006113 |
| EMISICD10\|F0021 | Dementia in Alzheimer's dis, atypical or mixed type, other symptoms, predominantly delusional | 1972251000006103 | 1972251000006119 |
| EMISICD10\|F0023 | Dementia in Alzheimer's dis, atypical or mixed type, other symptoms, predominantly depressive | 1972311000006108 | 1972311000006112 |
| EMISICD10\|F0000 | Dementia in Alzheimer's disease with early onset, without additional symptoms | 1971401000006107 | 1971401000006111 |
| EMISICD10\|F0013 | Dementia in Alzheimer's disease with late onset, other symptoms, predominantly depressive | 1972201000006102 | 1972201000006118 |
| EMISICD10\|F0093 | Dementia in Alzheimer's disease, unspecified, other symptoms, predominantly depressive | 1972451000006102 | 1972451000006118 |
| EMISICD10\|F0011 | Dementia in Alzheimer's disease with late onset, other symptoms, predominantly delusional | 1972181000006103 | 1972181000006119 |
| EMISICD10\|F0003 | Dementia in Alzheimer's disease with early onset, other symptoms, predominantly depressive | 1972131000006104 | 1972131000006115 |
| EMISICD10\|F0091 | Dementia in Alzheimer's disease, unspecified, other symptoms, predominantly delusional | 1972401000006101 | 1972401000006117 |
| EMISICD10\|F0012 | Dementia in Alzheimer's disease with late onset, other symptoms, predominantly hallucinatory | 1972191000006100 | 1972191000006116 |
| EMISICD10\|F0001 | Dementia in Alzheimer's disease with early onset, other symptoms, predominantly delusional | 1971541000006105 | 1971541000006114 |
| EMISICD10\|F0002 | Dementia in Alzheimer's disease with early onset, other symptoms, predominantly hallucinatory | 1971771000006108 | 1971771000006112 |

**Table S2.** The SNOMED codes and descriptions for T2D assignment.

| Original Read Code | Term | Snomed CT Concept Id | Snomed CT Description Id |
| --- | --- | --- | --- |
| L1809 | Gestational diabetes mellitus | 11687002 | 20191016 |
| C1001-2 | Non-insulin dependent diabetes mellitus | 44054006 | 493773010 |
| C10 | Diabetes mellitus | 73211009 | 121589010 |
| C10F | Type 2 diabetes mellitus | 44054006 | 197761014 |
| 1252 | FH: Diabetes mellitus | 160303001 | 249794016 |
| 1434 | H/O: diabetes mellitus | 161445009 | 251591016 |
| C10F-1 | Type II diabetes mellitus | 44054006 | 493774016 |
| C109-2 | Type 2 diabetes mellitus | 44054006 | 197761014 |
| 8CA41 | Diabetes mellitus diet education | 284350006 | 2575925018 |
| C109 | Non-insulin dependent diabetes mellitus | 44054006 | 493773010 |
| C108 | Insulin dependent diabetes mellitus | 73211009 | 121589010 |
| EMISNO3 | No H/O: diabetes mellitus | 857361000006103 | 857361000006119 |
| HNG0605 | [RFC] Diabetes mellitus | 908831000006102 | 908831000006118 |
| C10FJ | Insulin treated Type 2 diabetes mellitus | 237599002 | 2967820013 |

**Table S3.** The SNOMED codes and descriptions for migraine assignment.

| Original Read Code | Term | Snomed CT Concept Id | Snomed CT Description Id |
| --- | --- | --- | --- |
| F260-1 | Migraine with aura | 4473006 | 7595017 |
| F260 | Migraine with typical aura | 230462002 | 345348012 |
| F26 | Migraine | 37796009 | 63055014 |
| F261-1 | Migraine without aura | 56097005 | 93294013 |
| F261 | Common migraine | 56097005 | 93295014 |
| 1297 | FH: Migraine | 160342001 | 249890015 |
| 1474 | H/O: migraine | 161481007 | 251645017 |
| F261z | Common migraine NOS | 56097005 | 93295014 |
| F262 | Migraine variants | 193030005 | 297337016 |
| F262z | Migraine variant NOS | 193030005 | 297337016 |
| F26z | Migraine NOS | 37796009 | 63055014 |
| 8B6N | Migraine prophylaxis | 408381007 | 2159945013 |
| 14740 | History of migraine with aura | 608837004 | 2958210010 |
| EMISNQH/12 | H/O: migraine with aura | 1763401000006105 | 1763401000006114 |
| EMISNQH/13 | H/O: migraine without aura | 1763411000006108 | 1763411000006112 |
| ^ESCTCL257016 | Classical migraine | 4473006 | 7596016 |
| ^ESCTHI454016 | History of migraine | 161481007 | 2986612014 |
| ^ESCTCO476993 | Complex migraine | 193039006 | 2966546010 |
| ^ESCTMI500829 | Migraine aura without headache | 230465000 | 345351017 |
| ^ESCTRE701607 | Refractory migraine | 423894005 | 2644986016 |
| ^ESCTMI730629 | Migraine variant with headache | 445322004 | 2872940010 |
| ^ESCTMI752039 | Migraine with persistent visual aura | 699314009 | 2983774015 |
| ^ESCTCH803879 | Chronic intractable migraine without aura | 124171000119105 | 2967600014 |
| ^ESCT1234532 | Chronic migraine | 427419006 | 3638229012 |

**Table S4.** The blood borne measures and biometric data together with their averages and standard deviations. Data for these variables was collected for the case and control cohorts and only values falling within two standard deviations of the mean were included in the analysis.

| Measure | AVE | SD |
| --- | --- | --- |
| Neutrophil count | 4.581796 | 4.416166 |
| Basophil count | 0.04585 | 0.308122 |
| Platelet count | 267.4525 | 80.80724 |
| Monocyte count | 0.567595 | 1.788406 |
| Eosinophil count | 0.211243 | 0.881311 |
| Lymphocyte count | 2.168272 | 4.066914 |
| Serum C reactive protein level | 10.40646 | 24.78166 |
| Nucleated red blood cell count | 0.027757 | 0.463456 |
| Serum bilirubin level | 309.9447 | 305.2885 |
| Corrected serum calcium level | 2.383231 | 3.273162 |
| Serum high density lipoprotein cholesterol level | 1.457952 | 1.016703 |
| Serum low density lipoprotein cholesterol level | 2.885637 | 1.500549 |
| Plasma fasting glucose level | 5.666369 | 10.16407 |
| Serum albumin level | 42.00783 | 4.337708 |
| Serum sodium level | 139.6322 | 4.714234 |
| Serum aspartate aminotransferase level | 26.46625 | 26.42589 |
| Serum creatinine level | 83.31377 | 35.19167 |
| Serum urate level | 249.8492 | 190.4155 |
| Serum urea level | 5.977894 | 3.183229 |
| Serum luteinising hormone level | 13.79889 | 16.40006 |
| Plasma urea level | 5.744658 | 3.030966 |
| Serum thyroid stimulating hormone level | 2.540394 | 6.414843 |
| Plasma thyroid stimulating hormone level | 2.485047 | 7.091601 |
| HbA1c level | 7.257907 | 4.981173 |
| Serum free triiodothyronine level | 5.669793 | 3.804148 |
| Serum free T4 level | 15.01162 | 5.020864 |
| Serum total bilirubin level | 10.06324 | 8.484136 |
| Serum gamma-glutamyl transferase level | 54.16035 | 103.3814 |
| Plasma gamma-glutamyl transferase level | 50.21046 | 86.74896 |
| Plasma creatinine level | 83.43387 | 35.93568 |
| Plasma total bilirubin level | 11.00833 | 10.4315 |
| Plasma high density lipoprotein cholesterol level | 1.397647 | 0.419721 |
| Urine microalbumin level | 32.8716 | 109.1479 |
| Plasma albumin level | 40.66991 | 4.225833 |
| Plasma total cholesterol level | 4.952289 | 1.225716 |
| Plasma sodium level | 139.2439 | 3.313736 |
| Plasma alanine aminotransferase level | 26.62394 | 35.96136 |
| Plasma alkaline phosphatase level | 116.6087 | 84.45989 |
| Plasma calcium level | 2.375753 | 0.122061 |
| Plasma potassium level | 4.138521 | 0.950778 |
| Calculated low density lipoprotein cholesterol level | 2.833689 | 1.182399 |
| Serum creatine kinase level | 128.2204 | 126.8469 |
| Serum total cholesterol level | 5.028981 | 2.580017 |
| Urine albumin level | 35.93196 | 118.2024 |
| Urine creatinine level | 13.64487 | 90.30944 |
| Thyroid stimulating hormone level | 2.911864 | 11.787 |
| Total alkaline phosphatase level | 84.17497 | 50.05719 |
| Total white cell count | 7.417013 | 8.405416 |
| ALT/SGPT serum level | 28.48148 | 33.43329 |
| Blood glucose level | 6.572861 | 6.822651 |
| Haemoglobin A1c level | 7.717305 | 7.205028 |
| Serum glucose level | 5.765725 | 3.269726 |
| Serum follicle stimulating hormone level | 21.15659 | 29.55843 |
| Plasma glucose level | 5.932626 | 10.43315 |
| Serum ferritin level | 96.63587 | 142.6043 |
| Serum folate level | 8.56991 | 6.817203 |
| Serum globulin level | 28.67102 | 4.864038 |
| Serum inorganic phosphate level | 1.124378 | 1.289073 |
| Serum iron level | 14.83141 | 10.99456 |
| Serum potassium level | 4.428088 | 1.214448 |
| Serum testosterone level | 6.603195 | 8.670323 |
| Serum transferrin level | 3.252408 | 21.23239 |
| Serum triglycerides level | 1.665449 | 1.331858 |
| Serum vitamin B12 level | 403.5655 | 231.3958 |
| Serum alkaline phosphatase level | 87.00863 | 53.79808 |
| Serum bicarbonate level | 26.39662 | 4.142428 |
| Serum calcium level | 2.363842 | 1.642945 |
| Serum chloride level | 101.2795 | 9.543153 |
| Serum cholesterol level | 5.047981 | 2.307462 |
| Red blood cell count | 4.512244 | 4.189216 |
| Prostate-specific antigen level | 5.557065 | 36.24308 |
| Serum alanine aminotransferase level | 26.27311 | 33.63325 |
| Non high density lipoprotein cholesterol level | 3.469771 | 1.146355 |
| Haemoglobin A1c level | 46.1461 | 15.69581 |
| Calcium adjusted level | 2.381319 | 1.666903 |
| Serum prostate specific antigen level | 6.247608 | 41.4478 |
| Serum non high density lipoprotein cholesterol level | 3.427815 | 1.136847 |
| Serum total 25-hydroxy vitamin D level | 55.12612 | 28.85768 |
| Systolic arterial pressure | 133.3399 | 20.99563 |
| Diastolic arterial pressure | 78.29111 | 12.19942 |
| weight | 73.92255 | 26.27838 |
| height | 159.714 | 32.51027 |

